# Bringing accountability to the peak of the pandemic using linear response theory

**DOI:** 10.1101/2020.04.21.20068478

**Authors:** Meher K. Prakash

## Abstract

The peak of the daily new infections in COVID-19 remained qualitative in description and elusive in arrival. Because of the lack of clarity in what to expect from the peak, apart from the hope that one day the peak will be reached, there has been no metric to describe the success of the implemented strategies. We propose a way of predicting the number of infections that can be expected after a lockdown, assuming they come from the asymptomatic cases prior to the lockdown and using linear response theory. These predictions for several western countries faithfully follow the observed infections for several weeks after the lockdown, suggesting universalities in the recovery pattern of several countries. At the same time, the gap between the quantitative predictions of the recovery patterns for New York and Milan and the observations is striking. These gaps which arise even while emulating the recovery patterns of other western countries raise the possibility of an audit of the success of the implemented strategies, and the potential newer sources of infection.

## Introduction

The infections caused by the novel coronavirus (COVID-19) have exceeded 2 millions globally and continue to increase in numbers. The World Health Organization has declared COVID-19 as a pandemic, the first one in the 21^st^ century [1]. The concepts from the standard Susceptible-Infected-Recovered (SIR) model and its variants suggest that after a significant fraction of the susceptible individuals are infected, the spread of the infection slows down, resulting in a “peak” in between. However, even as little as 0.2% of infected population as of mid-April has already crippled the health care systems and economies of most countries. Hence tolerating more infections is not a sustainable option.

There are still many lacunae in the understanding of how this novel coronavirus spreads and who are susceptible to the infection [2,3]. There are presently no known drugs or vaccines for the COVID-19 pandemic and hence most governments resorted to strict nonpharmaceutical interventions [4] such as social distancing, and even lockdowns. Several epidemiological models [5-7] have been used as a way to guide these policy decisions [8]. The core principle guiding these strategies is the “flattening of the curve” [9] so that the number of infections become manageable by the available health care resources.

Models guided by the past intuitions from of the effects of active containment strategies such as the ones used for SARS by South Korea or COVID-19 by China, and seasonal variations on the virulence have predicted the “peak” of daily infections [10]. Most governments have imposed restrictions to halt the spread of the virus, albeit of varying levels of intensity and have been eagerly waiting to cross the peak, to mark the success of the containment strategies.

However, despite the qualitative picture of how a peak should look like, most seem to be clueless about when the peak will arrive, and if the trends they see in the numbers of new infections post lockdown may be considered satisfactory by their own standards or by the standards of other countries which implemented similar policies. To complicate this, there are stochastic fluctuations in the daily new case numbers. While universalities in the growth curves of different countries have been pointed out by matching the time when the different countries register 100 cases [10], a similar comparison across the countries for the effectiveness of the lockdown has been missing. To address some of these concerns which have been limiting the concepts to qualitative narrations of the peak, we introduce a model to quantitatively predict the new infections after the lockdown.

### Predicting the new daily infections after lockdown

In an infectious disease the infection spreads when an infected person, whether symptomatic or asymptomatic, comes in contact with susceptible individuals. It is now known that the reported COVID-19 infections are accompanied by a large number of asymptomatic infections [11]. After the lockdown, the first sign of success is a departure from the exponential growth behavior which suggests that the number of daily new infections (*I*(*t*)) does not depend on the number of persons identified as infected, but possibly on those who are exposed and still asymptomatic (*A*(*t*)). We adopt the convention that *t*=0 denotes the day of the lockdown. We model the new cases recorded after the lockdown as arising from the asymptomatic individuals *A*(-1), *A*(-2), *A*(-3).. with incubation times *t*+1,*t*+2,*t*+3, ..respectively. Thus the expression for the number of daily new infections *t* days after lockdown becomes

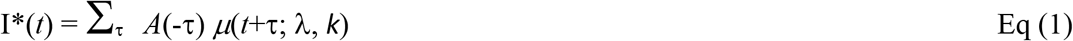

Where *μ*(*t*+τ; λ, *k*) represents the incubation time between the exposure and becoming symptomatic [12]. We conveniently assume this incubation time for COVID-19 to be a Weibull distribution defined by the parameters λ and *k*, as has been represented for many respiratory infections [13,14], although the conclusions will not change for other distributions. Eq. (1) which in physics is known as a linear response formalism [15], that has also been used for back-calculating the incubation time in the models for the spread of HIV [16]. We further assume that on a day τ days prior to the lockdown, the number of asymptomatic infections is proportional to the number of new infections recorded on that day (*A*(-τ) = *α*.*I*(-τ)), which modifies Eq. (1) to predict the daily new infections as

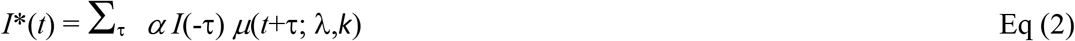

### Universality in the peak

We use the publicly available epidemiological data on the number of daily new COVID-19 infections [17] before the lockdown to predict the new cases after the lockdown. We first tested Eq. (2) by scanning over a range of parameters *λ, k* to find the best regression between the predictions (*I*\*(*t*)) and the observations (*I*(*t*)) predictions of Spain. As shown in **Figure 1A**, λ=16, *k*=2.0 gave a very good match with the observations up to a scaling factor *α*. It can be seen that the “peak” in the infections arises as a consequence of the peak in *μ*(*t*+τ; *λ, k*). Interestingly, we find that the same distribution function, *μ*(*t*+τ; λ=16, *k*=2), is also suitable for making predictions for Germany, Switzerland and Austria are shown in **Figures 1B-1D**. We call this incubation function which suits at least 4 different European countries as the Minimum European Incubation (MEI). This MEI shows that not just the growth curves from different countries are similar [10], but the relaxation following a lockdown implemented in a similar fashion is also similar. Of course, this similarity is not obvious unless one deconvolutes the history of infections each country went through until the lockdown to obtain the *μ*(*t*+τ; *λ, k*). Although incubation periods of 19 days [18] and 24 days [19] of COVID-19 infections have been reported, the median time has been estimated to be around 5.2 days based on the exposure history [20] of a few hundred individuals. The median time in the distribution we find common to the response of several countries (λ.(ln2)^1/k^=13.3 days), is clearly much larger. Whether the longer tails of the incubation have been missed out by the smaller sample sets of patient histories or whether the incubation period for the relaxation at the country level needs a different interpretation, merits a separate work. Here we use the MEI we derived for benchmarking the success of a lockdown.

**Figure 1.**
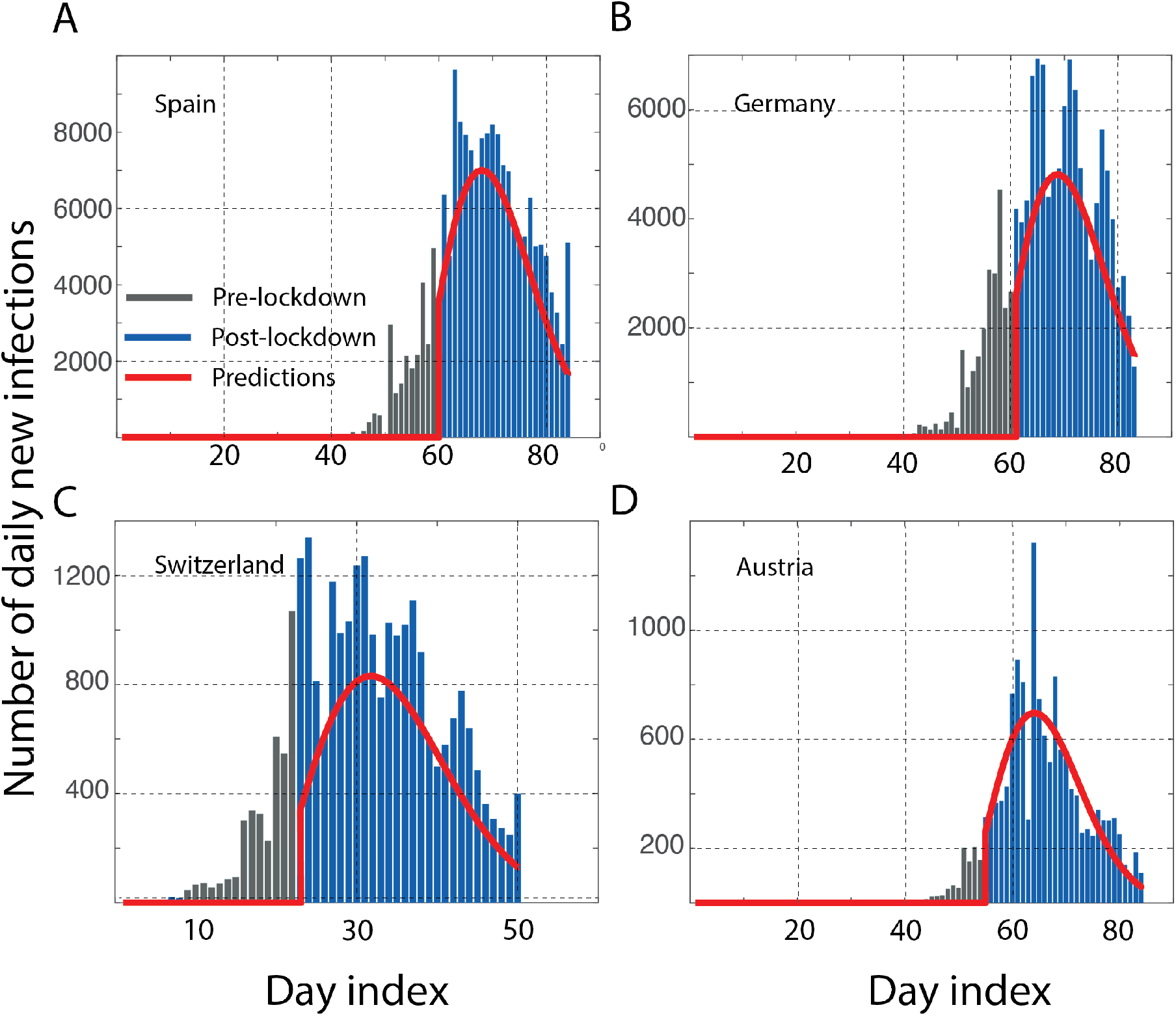
Universality of the relaxation patterns: The number of recorded daily infections before (gray bars) and after (blue bars) lockdown are shown along with the predictions based on Eq. 2. The data from four different European countries ***A***. Spain (lockdown, 14 March 2020) ***B***. Germany (lockdown, 21 March 2020) ***C***. Switzerland (lockdown, 17 March 2020) ***D***. Austria (lockdown, 16 March 2020) all show the best results with an incubation described by a Weibull distribution with parameters λ=16, *k*=2. The “peak” arises as a consequence of the peak in the incubation distribution. As with any stochastic process, the numbers do fluctuate around the fluctuations. The fact that all countries relax with similar *μ*(t+τ; *λ, k*) suggests a common Minimum European Incubation (MEI), as we refer to it for the relaxation. This “European standard” may be used as a benchmark for other countries with similar quality of quarantine measures. The data on the cases reported up to 15 April 2020 was obtained from the Johns Hopkins database.[17]

### Are the results of lockdown on track?

While different countries may use the same word lockdown to describe their mitigation efforts, the degree of restriction certainly varies. However, despite these differences, there might be a lowest common denominator which is set by several factors including the nature of the government as well as the cultural propensity for adherence to rules or a sense of rejection of the restrictions to personal liberty. One can see this commonality from the MEI that was noticed above for many European countries. We now use this as a benchmark metric to understand the evolution of the cases after their lockdown in two cities Milan and New York, assuming the biology of the virus and public response to lockdown are not significantly different. As one sees in Figure 2, the predictions made for Milan and New York with *μ*(*t*+τ; λ=16, *k*=2) clearly suggest a huge gap relative to the MEI. The differences in relaxation relative to an otherwise similar scenario from other western countries, or a purported peer-group, raises several natural questions - for any unknown reason these cities relax with a different incubation rate (λ=20, *k*=2; seems to suit with the present data)? However this argument is hard to support as the official norms suggest an extremely restricted movement in Milan compared even to its neighboring countries like Switzerland. Is the lockdown really as perfect as it is believed to be? Are there other sources for newer infections beyond just the asymptomatics before lockdown getting converted to infections? Clearly it cannot be through the individuals classified as infected, as that will reflect as an exponential growth all over again. Last but not the least, could these infections be because of the lack of the personal protective equipment, especially among the health care workers? While the benchmarks may be common, the differences can only be understood by a careful audit needs of the local realities.

**Figure 2.**
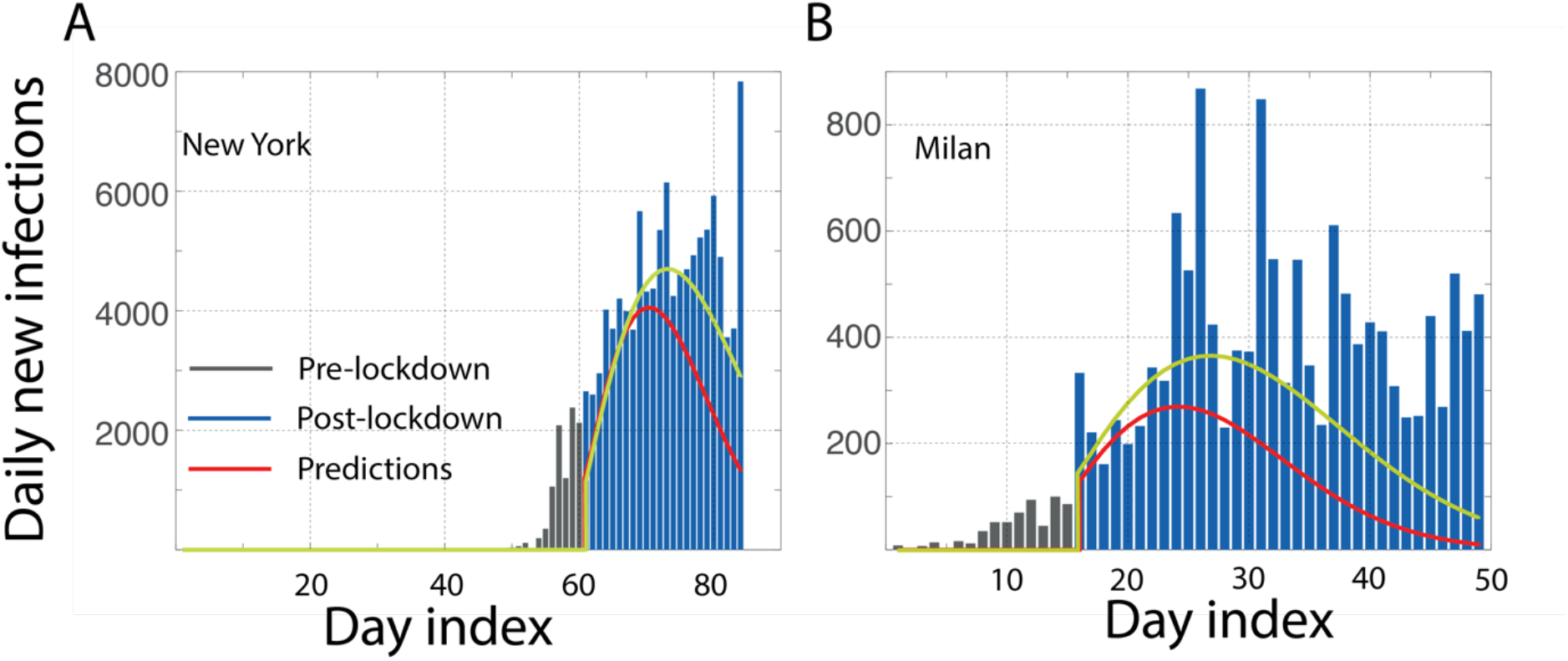
Judging the success of quarantine. We make predictions of daily new cases in two western cities ***A***. New York and ***B***. Milan, assuming the biology of the disease and the quality of quarantine to be similar. Using the MEI incubation time described by *μ*(t+τ; λ=16, k=2), we see a very large gap between the predictions (red line) and the observations for both the cities. Predictions (yellow line) made with *μ*(t; λ=20, *k*=2) are also shown for a reference, which also are still not satisfactory. These gaps raise questions which are beyond the obsessive “did we reach a peak” question, such as why is the relaxation so different for these cities? Are there other sources of infection beyond the asymptomatics converting into symptomatics.

## Conclusions

In conclusion, we introduce a way of predicting the daily new infections after a lock-down. This framework allows one to transition from a very naïve desire of seeking a peak to actively tracking the progress on a daily basis, ranking themselves among the peers with comparable governments and cultural backgrounds and possibly identifying other sources of infection that were not even suspected earlier when there was no quantitative expectation from the “peak”.

## Data Availability

The analyses have been performed from publicly available data. However, upon request the specific analysis folders including the data and scripts will be provided.

## Acknowledgements

We thank Prof. Shobhana Narasimhan for helpful comments.

